# Caregiver differentiation between dystonia and spasticity in cerebral palsy

**DOI:** 10.64898/2026.02.24.26347000

**Authors:** Alyssa Rust, Emma Lott, Susie Kim, Michele Shusterman, Lilly Shusterman, Danielle Guez-Barber, Fayza Jaleel, Amy McQueen, Bhooma Aravamuthan

## Abstract

**Background:** Dystonia is a debilitating movement disorder that is difficult to assess when co-existing with spasticity, as is typical in cerebral palsy (CP). Querying caregivers about their children’s movements is known to increase clinical dystonia identification. However, beyond identification, determining whether dystonia is the predominant vs. accompanying movement feature in a child with CP can guide clinical decision making, particularly regarding surgical candidacy.

**Objective:** To determine whether caregivers’ movement descriptions differed between children with predominant dystonia, predominant spasticity with accompanying dystonia, and predominant spasticity without dystonia.

**Methods:** In this cross-sectional study, we used conventional content analysis to codify caregivers’ descriptions of triggered involuntary movements in children with CP seen in a tertiary care CP center between 4/2023 and 12/2024. Movement feature frequencies were compared across tone types using Chi-square tests with Bonferroni corrections for multiple comparisons.

**Results:** Of 180 children with CP (mean age 9.2, 47.8% male), caregivers of children with predominant dystonia (50/180, 27.8%) more frequently described movements triggered by negative emotions (p<0.002) and affecting their back, trunk, and whole body (p<0.04). Caregivers of children with predominant spasticity with dystonia (99/180, 55.0%) more frequently described movements affecting a single limb (p<0.04). Caregivers of children without dystonia (31/180, 17.2%) described movements as being slight or small (p<0.008). These differences persisted even for caregivers unaware their child had dystonia (77/149, 51.6%).

**Conclusions:** Caregivers’ movement descriptions differ between children with different combinations of dystonia and spasticity, which may help inform clinical management and guide communication with families about dystonia.

## Introduction

Dystonia is a painful movement disorder that can significantly limit motor function and is notoriously difficult to diagnose.(1–4) This is particularly true when dystonia co-exists with spasticity, which requires a distinct treatment regimen.(5–7) The presence of dystonia can rule in or rule out many pharmacologic and surgical treatment options, particularly in people with cerebral palsy (CP) who commonly have co-existing dystonia and spasticity.(5,6,8,9) CP is the most common lifelong motor disability and the most common condition associated with dystonia in childhood.(10–13) Approximately 15% of people with CP have dystonia as their predominant movement/tone type as defined by the Surveillance of Cerebral Palsy in Europe (SCPE) classification system.(14,15) However, at least 70% of people with CP have less severe dystonia that accompanies spasticity as their predominant tone type.(16) Differentiating between whether someone has predominant dystonia vs. dystonia accompanying predominant spasticity vs. no dystonia at all may help guide treatment decisions more readily than simply identifying the presence or absence of dystonia alone. Though dystonia is often considered a contra-indication to selective dorsal rhizotomy and orthopedic procedures used to treat spasticity and its musculoskeletal sequelae in children with CP, a child whose functional limitations are due predominantly to spasticity, and not their accompanying dystonia, may still benefit from these procedures.(17–19) Therefore, it is important not just to identify dystonia, but to determine the degree to which it is present in a person with CP.

Because dystonia often variably appears in the same individual, we have shown that repeated longitudinal evaluation is valuable for identifying dystonia.(3) The person with the greatest longitudinal exposure to a child with CP is their caregiver. We have shown that caregivers can recognize dystonic movements in their child.(20,21) Compared to caregivers of children with CP without dystonia, caregivers of children with CP and dystonia are more likely to report that their children have involuntary movements triggered (i.e. brought out or worsened) by voluntary movement or another person handling them,(20) in line with consensus definitions for dystonia(22,23) and screening exam maneuvers for identifying dystonia in the clinician-facing Hypertonia Assessment Tool.(24) Similar to increased rates of autism diagnosis after implementation of caregiver-facing screeners like the Modified Checklist for Autism in Toddlers (M-CHAT),(25) we have shown that incorporating caregiver-facing questions about tone variability into clinical dystonia screening can increase rates of dystonia diagnosis in children with CP.(21) However, it remains unclear whether caregivers’ descriptions of their children’s movements can move beyond identification of dystonia and additionally help distinguish between predominant dystonia, dystonia accompanying predominant spasticity, and absence of dystonia in children with CP. Having caregivers contribute to this differentiation could facilitate clinical management decisions, particularly regarding surgical candidacy for children with CP.(17–19) In this cross-sectional study, we compare caregivers’ descriptions of their children’s movements in those with A) predominant dystonia, B) predominant spasticity with accompanying dystonia, and C) predominant spasticity without dystonia. We hypothesize that caregivers’ descriptions of their children’s distinct movement patterns will differ between these three mutually-exclusive groupings, information that may ultimately be valuable for developing caregiver-facing dystonia assessments, increasing caregiver and family awareness of how dystonia presents in children with CP, and facilitating clinical management decisions.

## Methods

### Standard Protocol Approvals, Registrations, and Patient Consents

Human Subjects Research exemption was granted by the Washington University Institutional Review Board (ID#202309003, 09/11/2023).

### Respondents and data collection

Data was collected from caregivers of children with CP during routine clinical care at the St. Louis Children’s Hospital Cerebral Palsy and Mobility Center as a part of our standardized intake assessment emailed to families as a REDCap link (see bit.ly/CP-Intake-Methodology).(9) As a part of this intake, we asked caregivers two questions about triggered involuntary movements that we have previously shown prompt caregivers to identify potentially dystonic movements in their child.(20) Caregivers that respond ‘yes’ to either question (indicating that they have observed triggered involuntary movements in their child) are then prompted to enter a text description of these movements. We additionally asked caregivers their knowledge of their child’s tone and movement disorders diagnoses (Figure 1).

**Figure 1.**
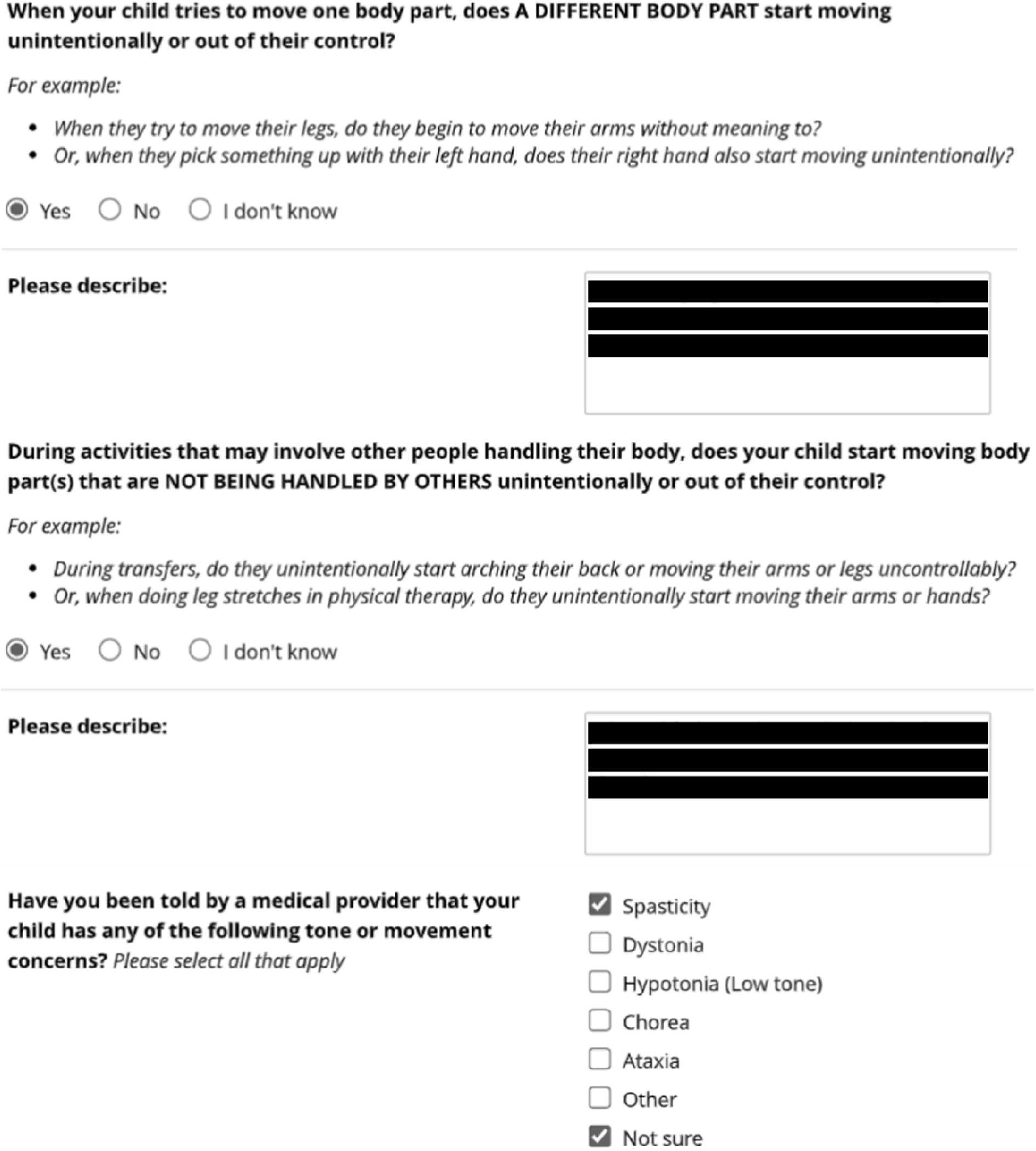
Questions posed to caregivers regarding triggered involuntary movements and their knowledge of their child’s tone diagnoses.

Separately, CP Center clinicians (pediatric movement disorders physicians specializing in CP) were asked to identify the child’s predominant tone type as described in the SCPE classification system(14,26) and whether dystonia was present in the child using the validated Hypertonia Assessment Tool.(24) This information is entered by the clinician into the medical record using drop down menu selections as a part of every single CP Center visit during routine clinical care (see bit.ly/CP-Intake-Methodology).(9)

Inclusion criteria were people seen in our CP Center between 4/3/2023 and 12/18/2024 whose caregivers had observed triggered involuntary movements in their child and who were identified by a CP Center clinician as either having predominant dystonia, predominant spasticity with accompanying dystonia, or predominant spasticity without dystonia. The CP diagnosis was established based on CP Center clinician expertise as documented explicitly by them in the medical record guided by the 2006 CP definition(27) and literature operationalizing this definition for clinical application.(28,29) Exclusion criteria were people with CP whose caregivers did not provide written descriptions of the triggered involuntary movements they observed in their children.

### Qualitative analysis

Caregivers’ movement descriptions were analyzed using conventional content analysis.(30) To fully characterize response content, we built upon our established code book generated in our previous work analyzing caregivers’ movement descriptions.(20) Two investigators (AR, EL) independently coded all caregivers’ movement descriptions with discrepancies resolved in consensus with additional input as necessary from a third independent coder (BRA).

### Quantitative and statistical analysis

We separated our subjects into three groups for analysis: those with predominant dystonia, predominant spasticity with dystonia, and predominant spasticity without dystonia. Notably, the vast majority of children with CP and predominant dystonia also have accompanying spasticity,(9) precluding meaningful differentiation of children with predominant dystonia into those with and without accompanying spasticity. Only codes (i.e. movement features) cited at least 5 times were used for quantitative analysis to allow for differentiation in code frequency between three groups. Code frequencies were compared between groups using Chi-square tests with Bonferroni corrections for multiple comparisons if expected code frequencies were greater than 5. Fisher’s exact tests with Bonferroni corrections were used if expected frequencies were less than 5. Significance levels were set a priori at *p*<0.05. Statistical analyses were performed using GraphPad Prism (version 8, GraphPad Software).

### Data availability

De-identified data will be shared with investigators upon reasonable request.

## Results

### Respondent demographics

We identified 234 subjects meeting inclusion criteria of whom 54 (23.1%) were excluded because caregivers did not provide written descriptions of the triggered involuntary movements they observed. This yielded 180 subjects for analysis (average 9.2 years old, ranging from 6 months to 23 years old, 95% CI 8.4-9.9 years, 86/180 or 47.8% male, 84/180 or 46.7% independently ambulatory). Of these 180 people, 50 (27.8%) had predominant dystonia, 99 (55.0%) had predominant spasticity with dystonia, and 31 (17.2%) had predominant spasticity without dystonia. As expected, most people with predominant dystonia had accompanying spasticity (42/50, 82%). Of children diagnosed with dystonia, the majority (77/149, 51.7%) had caregivers who were unaware of this diagnosis, including caregivers of children with predominant dystonia (27/50, 34%).

### Movement features cited by caregivers

Caregivers described 339 triggered involuntary movements with movement features that we found fell into five different code categories: trigger descriptor (n=233), trigger body region (n=104), action descriptor (n=304), action body region (n=293), and severity/frequency descriptor (n=96). Example quotes and coding across these categories are shown in Table 1. All subcodes cited at least 5 times are shown divided by code category in Figures 2-3.

**Table 1.** Example caregiver movement description quotes and corresponding movement feature codes.

| Example quotes | Movement features |  |  |  |  |
| --- | --- | --- | --- | --- | --- |
|  | Trigger descriptor | Trigger region body | Action descriptor | Action body region | Severity/frequency descriptor |
| "When he is trying to do a task with righty (his affected hand) typically you see lefty trying to do the same." | fine motor task | hand (affected) | mimic | hand (contralateral) | typical |
| "his arms and legs all stiffen when he is active, for the most part." | move |  | stiffen | arms and legs (bilateral) | typical |
| "If grasping with one hand the opposite arm stiffens outward." | grasp | hand (unilateral) | stiffen | arm (contralateral) |  |
| "Sometimes when playing with toys with his right hand his left hand does not stay still" | fine motor task | hand (unilateral) | move | hand (contralateral) | sometimes |
| "When excited or moving/running, left arm tightens and raises" | excited (emotion-positive); move; run |  | stiffen; raise | arm (unilateral) |  |
| "she will move her right arm and her left leg will move" | move | arm (unilateral) | move | leg (contralateral) |  |
| "Right side moves uncontrollably when excited" | excited |  | move | body (unilateral) |  |
| "when the right arm goes up, the left arm normally follows a little." | raise | arm (unilateral) | mimic | arm (contralateral) | typical; slight/small |
| "sometimes if we take her arm and try to help her grab something her other arm will move" | handle | arm (unilateral) | move | arm (contralateral) | sometimes |
| "back arches unintentionally when doing most things" | move |  | arch | back | typical |
| "She usually tenses her body up whenever it's something she doesn't like or want to do." | frustration (emotion-negative) |  | stiffen | body | typical |
| "arching back when being moved" | transfer |  | arch | back |  |
| "When transferring his legs extend" | transfer |  | extend | legs (bilateral) |  |
| "Strong extension pattern backward" |  |  | extend | body | strong |
| "He is always moving his body." |  |  | move | body | always |
| "He has a hard time controlling any other muscles but the ones he is currently using." | move |  | overflow |  |  |

**Figure 2.**
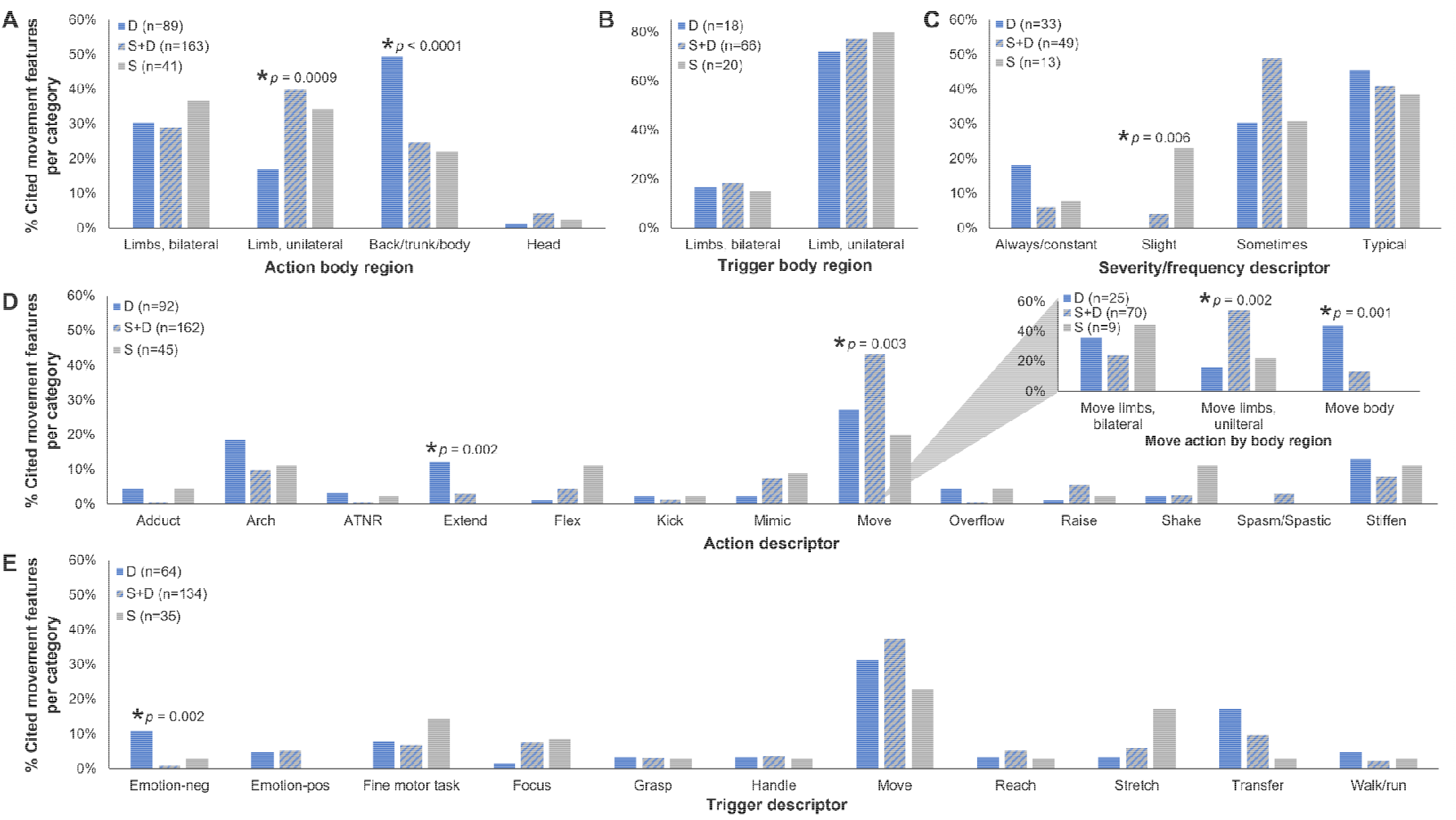
Movement features cited by caregivers across feature categories and tone types. Movement features cited by caregivers were compared between children with predominant dystonia (D – blue), predominant spasticity with accompanying dystonia (S+D – blue/grey stripe), and predominant spasticity without dystonia (S – grey) within a given feature category: action body region (A), trigger body region (B), severity/frequency descriptor (C), action descriptor (D), and trigger descriptor (E). Indicated N’s are for the number of times caregivers of children with each tone type cited features within the indicated feature category. Comparisons are by Chi-square test with post-hoc comparison of proportions with Bonferroni corrections for multiple comparisons (*p<0.05).

**Figure 3.**
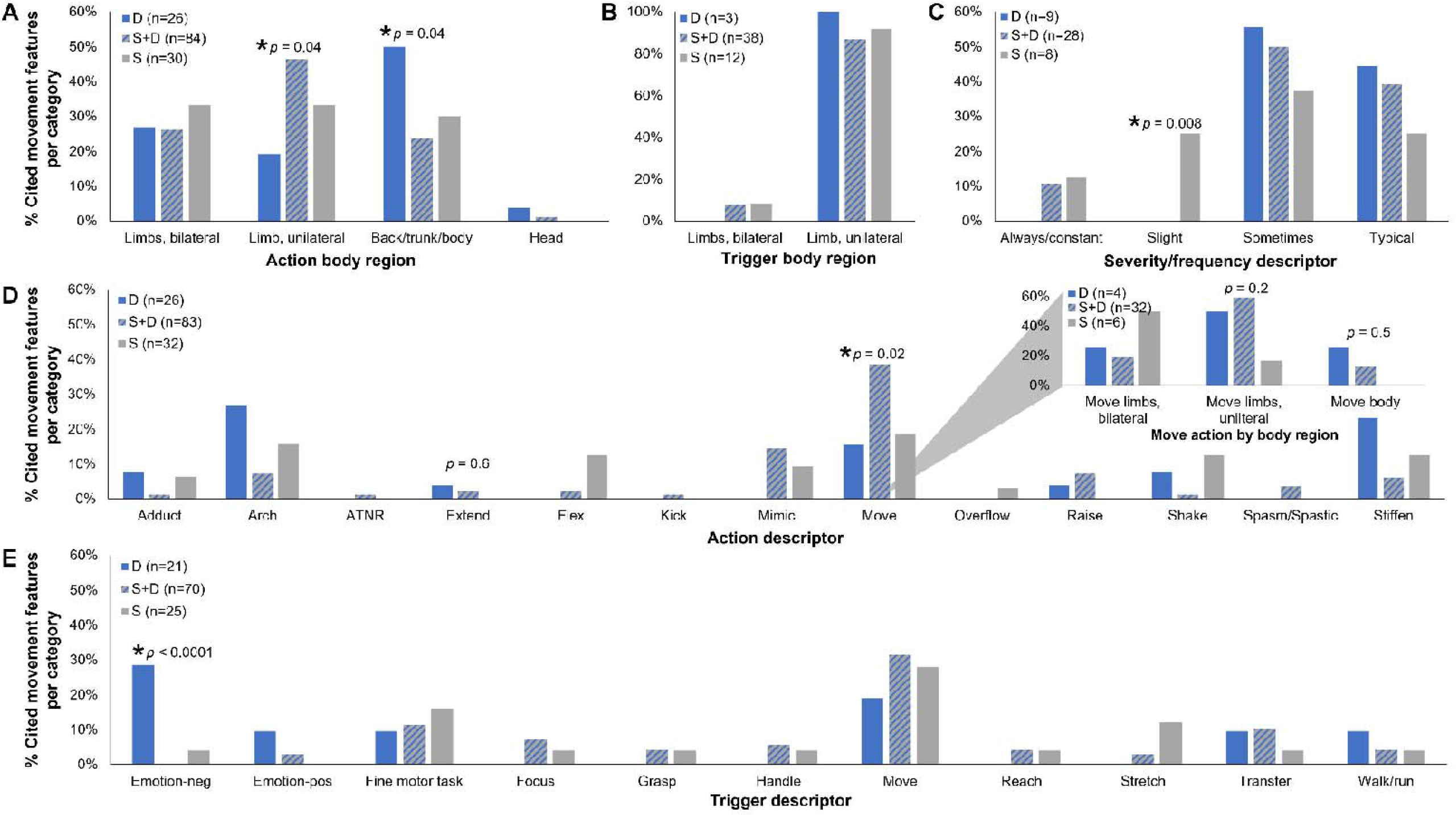
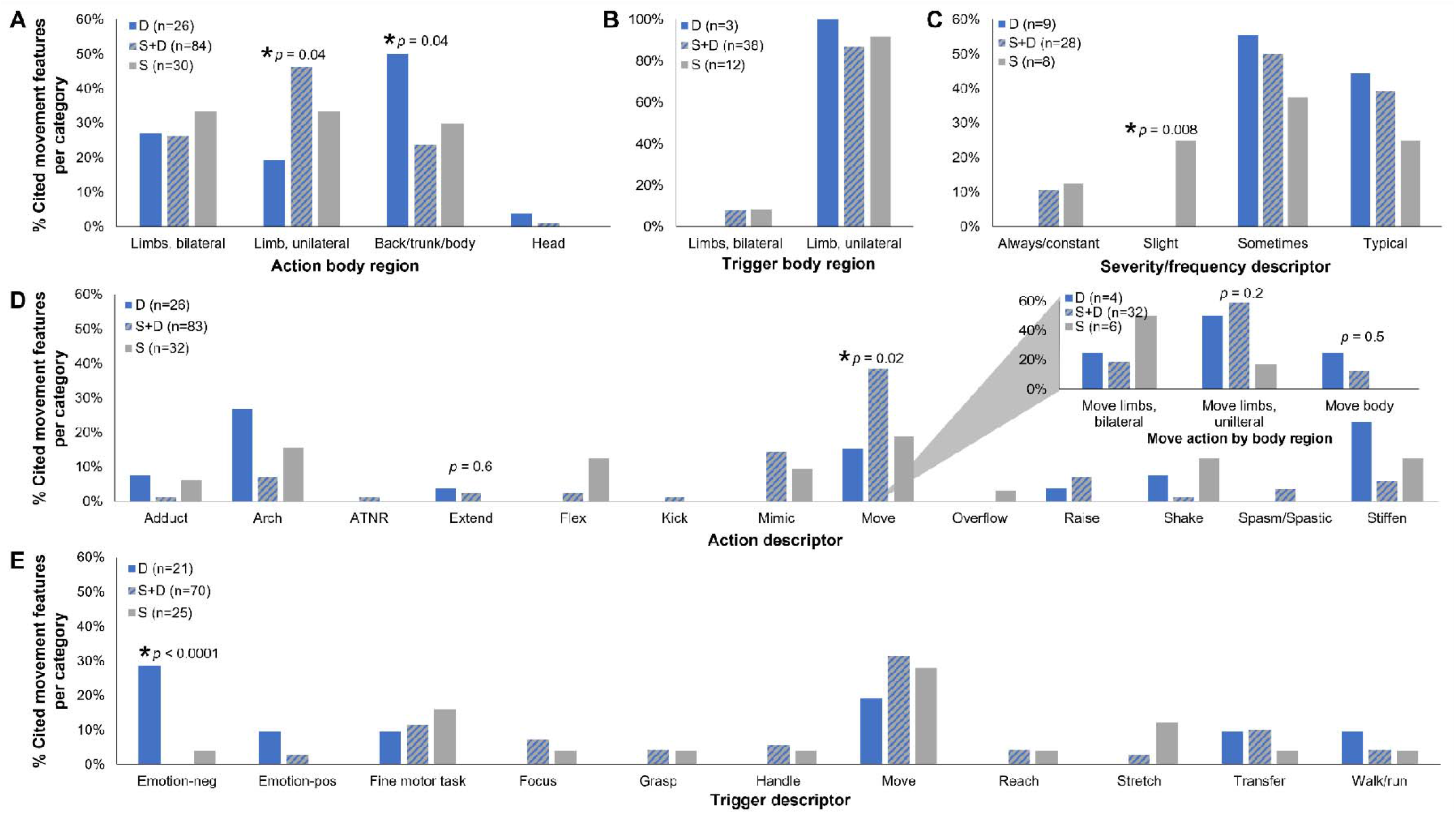
Movement features cited by caregivers who did not indicate their child was diagnosed with dystonia across feature categories and tone types. Movement features cited by caregivers were compared between children clinically diagnosed with predominant dystonia (D – blue), predominant spasticity with accompanying dystonia (S+D – blue/grey stripe), and predominant spasticity without dystonia (S – grey) within a given feature category: action body region (A), trigger body region (B), severity/frequency descriptor (C), action descriptor (D), and trigger descriptor (E). Indicated N’s are for the number of times caregivers of children with each tone type cited features within the indicated feature category. Comparisons are by Chi-square test with post-hoc comparison of proportions with Bonferroni corrections for multiple comparisons (*p<0.05).

Caregivers described movements more commonly affecting the back, trunk, or whole body in people with predominant dystonia (42/89, 47.2%) compared to people with predominant spasticity either with (34/163, 20.9%) or without dystonia (9/41, 22.0%) (p<0.0001). In contrast, people with predominant spasticity had movements described as more commonly affecting a single limb, regardless of whether they additionally had dystonia (65/163, 39.9%) or not (14/41, 34.1%), compared to those with predominant dystonia (15/89, 16.9%) (p=0.0009) (Figure 2A). Caregivers more frequently described movements as slight or small in people without dystonia (3/13, 23.1%) compared to those with predominant spasticity and dystonia (2/49, 4.1%) or predominant dystonia (0/33, 0.0%) (p=0.006) (Figure 2C).

Describing the action generically as movement (as opposed to more specific terms like stiffening, raising, or arching) was more frequent for caregivers of people with predominant spasticity with dystonia (70/162, 43.2%) compared to people without dystonia (9/45, 20.0%) or predominant dystonia (25/92, 27.2%) (p=0.003) (Figure 2D). However, this was true only for generic descriptions of movement of a single limb (p=0.002). Descriptions of generically moving the whole body were still most common in people with predominant dystonia (p=0.0001) (Figure 2D, inset). Caregivers of people with predominant dystonia also more frequently described extension movements (11/92, 12.0%) compared to those with predominant spasticity either with (5/162, 3.1%) or without dystonia (0/45, 0.0%) (p=0.002, Figure 2D). Finally, caregivers of people with predominant dystonia more frequently described involuntary movements triggered by negative-valence emotions (e.g. fear or frustration as opposed to a positive-valence emotion like excitement) (7/64, 10.9%) compared to those with predominant spasticity either with (1/134, 0.7%) or without dystonia (1/35, 2.9%) (p=0.002, Figure 2E).

### Movement features cited by caregivers unaware their child was diagnosed with dystonia

Caregivers of 102 people reported that the person they cared for had not been diagnosed with dystonia. Of these 102 people, 17 (16.7%) had been identified by a clinician to have predominant dystonia, 60 (58.8%) had predominant spasticity with dystonia, and 25 (24.5%) had predominant spasticity without dystonia.

Even caregivers who were unaware their child had dystonia cited many of the same movement features at comparable frequencies across children with different tone types. Involuntary movements of the back, trunk, and whole body (p=0.04, Figure 3A) and negative valence emotions as a trigger (p<0.0001, Figure 3E) were cited most frequently for children with predominant dystonia. Movements affecting a single limb were cited most frequently for children with predominant spasticity with dystonia (p=0.04, Figures 3A). Slight or small movements were cited most frequently by caregivers of children without dystonia (p=0.008, Figure 3C).

## Discussion

Compared to caregivers of people with predominant spasticity, caregivers of people with predominant dystonia more frequently describe their triggered involuntary movements to involve their back, trunk, and whole body and be triggered by negative valence emotions. In contrast, caregivers of people with predominant spasticity with dystonia more frequently describe their children having triggered involuntary movements that affect a single limb. Caregivers of people without dystonia describe triggered involuntary movements as being slight or small. Importantly, these descriptions are not dependent on the caregiver knowing that their child has been diagnosed with dystonia. These findings support the vast expertise of caregivers in supporting motor phenotyping of children with CP.

We have previously demonstrated that asking caregivers about triggered involuntary movements can facilitate clinical identification of dystonia.(20,21) Our results additionally suggest that asking caregivers about the regions of the body affected by triggered involuntary movements, the types of triggers, and the perceived size or amplitude of the movement may help differentiate between predominant dystonia, dystonia accompanying predominant spasticity, and predominant spasticity without dystonia. This differentiation can guide clinical management particularly when considering surgical interventions targeted to spasticity and its sequelae.(17–19)

Determination of dystonia as a predominant vs. accompanying feature of CP may give a crude approximation of dystonia severity. Determination of the predominant movement/tone type in CP includes consideration of the number of limbs/body regions affected (14,26) and most dystonia severity scales sum dystonia severity across all body regions to generate an aggregate severity score.(31–33) Noting this potential association, it is reassuring that the movement features caregivers recognize in their children with dystonia are already included in clinician-facing dystonia severity scales.(31–33) Future work could incorporate the salient movement features identified in this study to develop a detailed caregiver-facing assessment for dystonia in CP.

Of the children in the study population diagnosed with dystonia, approximately half had caregivers who were unaware of this diagnosis, even when dystonia was the predominant tone type. This finding alone is remarkable: caregivers are often unaware of dystonia diagnoses made by their child’s clinician. Even if clinicians are regularly sharing information about dystonia with families, our results suggest clinicians are not communicating this information in a way that families understand and/or retain. Noting that “improving family awareness of dystonia” is a key part of a collaboratively-set research agenda for dystonia in CP,(34) our results suggest language that can be used to describe dystonia to caregivers and families. Family-facing educational materials on how dystonia may present in children with CP, like the Dystonia in CP Toolkit recently published by the Cerebral Palsy Research Network,(35) may additionally help address this knowledge gap.

### Limitation and future directions

Clinician assessment was at a single timepoint, which is an important limitation noting that dystonia identification improves with longitudinal assessment.(3) Additional descriptions collected from multiple centers and geographical regions, including from centers that are not CP centers, would allow for a broader characterization of the language caregivers use to describe their children’s movements.

Noting that some movement features that were cited relatively infrequently differed between groups, future studies should query caregivers about these phenomena explicitly and/or use a larger pool of caregivers to confirm the reproducibility of the results presented here. These future studies can further explore caregivers’ movement descriptions including the types and intensity of negative-valence emotions that may act as dystonia triggers.

Caregivers provided written descriptions of their children’s triggered involuntary movements after reading prompts that included general examples of such movements (Figure 1). These examples could have influenced the content of caregivers’ descriptions, though caregivers also described many movements and triggers that were not explicitly mentioned in the prompts. Notably, all caregivers received the same set of prompts and still described their children’s triggered involuntary movements significantly differently between groups.

We focused this study on differentiation of dystonia from spasticity, by far the most common movement conditions seen in people with CP.(8,9) At our CP Center, as is done in 50% of CP registries globally,(36) hypotonia is considered to be a predominant tone type in people with CP. Though we have recently argued for the inclusion of hypotonia as a CP tone type,(29) our work also shows this is an area of significant clinical variability.(28,37) Though people with predominant hypotonia were excluded from this analysis, future work can additionally probe caregiver descriptions of hypotonia. In addition, the much rarer phenomena of ataxia, chorea and athetosis (present in less than 3% of people with CP as we have previously published(9)) should also be probed in future work.

It is likely that the movements described by caregivers vary by age of the person they care for. Future work should prioritize the necessary sample sizes to stratify people with CP by age and include caregivers of older individuals. Future work should also analyze how older children and adults describe their own movements, which would illuminate how people directly affected by dystonia experience it themselves.

## Conclusions

Caregiver descriptions of triggered involuntary movements in people with CP differ significantly between people with predominant dystonia, predominant spasticity with accompanying dystonia, or predominant spasticity without dystonia. These results suggest that caregivers could help differentiate between these CP tone patterns, which could facilitate clinical management decisions particularly regarding surgical candidacy. These results additionally suggest wording that can be used to explain dystonia to families and may lay the groundwork for developing caregiver-facing assessments for dystonia in CP.

## Acknowledgement

We gratefully acknowledge all our participating families for sharing their unique expertise on their children’s movements. We are proud to celebrate their expertise in this study.

## Authors’ roles

Dr. Aravamuthan was responsible for the design, execution, analysis, writing, and editing of final version of the manuscript.

Dr. Rust and Ms. Lott were responsible for the execution, analysis, and editing of final version of the manuscript.

Ms. Kim was responsible for execution and editing of final version of the manuscript.

Ms. Michele Shusterman, Ms. Lily Shusterman, Dr. Barber, Ms. Jaleel and Dr. McQueen were responsible for analysis and editing of the final version of the manuscript.

All authors approved the final manuscript as submitted and agree to be accountable for all aspects of the work.

